# Adherence and impact of the World Health Organization zinc and antibiotic administration guidelines for diarrhea management among children aged 6-35 months: EFGH study, 2022 - 2024

**DOI:** 10.64898/2026.02.10.26346032

**Authors:** Adama Mamby Keita, Erika Feutz, Milagritos D. Tapia, Patricia B. Pavlinac, Kirkby D Tickell, Alex O. Awuor, Rebecca Oketch, Samba Sow, Karen L. Kotloff, M. Jahangir Hossain, Jen Cornick, Nigel A. Cunliffe, Margaret N. Kosek, Maribel Paredes_Olortegui, Firdausi Qadri, Farhana Khanam, Farah Naz Qamar, Muhammad Tahir Yousafzai, Elizabeth Rogawski McQuade

## Abstract

**Background:** Diarrheal disease is the second leading cause of under-five mortality and morbidity in Sub-Saharan Africa. The World Health Organization (WHO) has developed treatment guidelines to support clinicians in the management of pediatric diarrhea; however, adherence to, and the impact of, these guidelines are not well described in low- and middle-income countries.

**Method:** We conducted a secondary analysis of data from the Enterics for Global Health study to determine whether adherence to WHO diarrhea management guidelines, specifically zinc supplementation and antibiotic administration (when appropriate), shortened the duration of diarrhea among children aged 6-35 months who presented to selected health facilities with diarrhea.

**Results:** This analysis includes 9,397 children aged 6 to 35 months with diarrhea enrolled across all seven EFGH sites. The majority (63.3%) of participants were under 18 months of age and 54.4% were male; 1,214 children (12.9%) presented to care with dysentery. Zinc was frequently administered (96.6%), with over 10 days of mean prescribed treatment duration. Of 5,061 children (53.9%) offered antibiotics, 3,082 (60.9%) received a WHO recommended regimen. Among participants who presented with dysentery, 67.6% were prescribed a WHO-recommended antibiotic. Among participants with watery diarrhea without dysentery, 72.4% were not prescribed any of the recommended antibiotics and were thus considered guideline-adherent. Overall, 6,302 (67.1%) children received guideline-adherent care when combining the zinc and appropriate antibiotic use indicators. In children with dysentery, providing WHO-recommended antibiotics was associated with 1.08 (95% CI: 0.53, 1.53) fewer days of diarrhea than those with dysentery who did not receive antibiotics. Children who were given 10+ days of zinc had on average 0.36 (95% CI: 0.03, 0.70) fewer days of diarrhea.

**Conclusion:** We found that two-thirds of children in this study received guideline adherent care in terms of zinc and appropriate antibiotic use for the treatment of childhood diarrhea, and that adherent treatment was associated with shorter duration diarrhea.

## Introduction

Diarrheal diseases are the second leading cause of under-five mortality and morbidity in Sub-Saharan Africa (1). In 2003, about 1.87 million children aged less than five years died from diarrhea, with 80% of those deaths occurring in the first year of life. On average, children under three years of age in low-and middle-income countries (LMICs) experience about three episodes of diarrhea every year (2), making appropriate management of diarrheal illness critical in these settings. Zinc plays a role in cell growth as a micronutrient and antioxidant, thereby strengthening the immune system and reducing the duration and severity of diarrheal episodes; there is evidence that zinc supplementation also prevents future diarrheal episodes (3). Antibiotics can be beneficial for the treatment of bloody diarrhea, which is often caused by bacteria and parasites(4).

The World Health Organization (WHO), in coordination with regional health ministries, have developed treatment guidelines to support clinicians in the management of pediatric diarrhea. Based on randomized controlled trial evidence of benefits (5,6), WHO recommends rehydration for dehydrated children, antibiotics for suspected dysentery or cholera, and therapeutic zinc to reduce diarrhea duration and replenish zinc stores (2,4). Adherence to WHO guidelines varies from low to suboptimal in different countries in the world (7,8). In Mali, among the Vaccine Impact on Diarrhea in Africa (VIDA) study participants, approximately 92.3% children with diarrhea and severe dehydration did not receive zinc; 79.5% did not receive IV rehydration and 96.6% did not receive oral rehydration solution (ORS) (7). In South Africa, a study on the management of acute diarrheal diseases reported that 84% of patients did not receive appropriate antibiotics (9). The implications of such suboptimal adherence include increasing mortality and antimicrobial resistance (AMR), the latter a result of inappropriate antibiotic use for non-bacterial diarrheal illness (8,10).

Among under-five children with diarrheal illness in LMICs, familial financial limitations, concurrent fever, and use of unauthorized drug vendors were identified as factors affecting the adherence to, and effectiveness of, treatment (11). In 2014, in a cross-sectional study in Lagos, Nigeria, Okafor et al found that mothers had poor knowledge of oral zinc and its role in diarrhea management (12). Education, among both healthcare providers (to reinforce existing guidelines) and members of the community is likely to increase guideline adherence for diarrheal illness (13,14).

Documenting and characterizing risk factors for sub-optimal adherence to WHO guidelines can guide areas for quality improvement. In particular the association of guideline adherence to important health outcomes, such as duration of diarrhea and healthcare utilization (and associated increase in costs), may provide important evidence for policy change in regions with high non-adherence rates. In a secondary analysis of the Enterics for Global Health (EFGH) *Shigella* surveillance study, we described the characteristics of children receiving WHO guideline-adherent clinical management for diarrhea and determined whether adherence to WHO diarrhea management guidelines (zinc and appropriate antibiotics for dysentery and cholera) shortened the duration of diarrhea among children aged 6-35 months who presented to selected health facilities with diarrhea.

By conducting this study, we aim to provide regional data in support of quality improvement initiatives to optimize the management of diarrheal illness.

## Methods

### Study design

As described previously, EFGH study is a 2-year, multi-center, facility-based surveillance study which aims to gather data to identify potential sites for Phase IIb and Phase III *Shigella* vaccine trials across select health facilities in Bangladesh, Kenya, Malawi, Mali, Pakistan, Peru, and The Gambia. The primary aim of the study was to determine the incidence of *Shigella*-attributed medically-attended diarrhea (MAD) in children 6 to 35 months of age at each site (15).

We performed a secondary analysis of data from the diarrhea case surveillance component of EFGH to determine whether adherence to WHO guidelines (zinc and antibiotic therapy, when appropriate) shortened the duration of diarrhea among children aged 6-35 months presenting to selected health facilities.

### Study population

The study population is children aged 6-35 months in all included sites meeting criteria for EFGH enrollment (presenting to health facilities with diarrhea, dysentery, or gastroenteritis) with complete records.

### Data collection

Demographic information about each participant, including sex, age, comorbidities, and family history, was collected upon the child’s enrollment into EFGH using standardized caregiver and clinician questionnaires. Diarrhea management, including zinc and antibiotic administration, was recorded by study staff throughout the child’s enrollment visit at the health facility. Treatments administered at the health facility and those prescribed to the child for home use were included to define guideline adherence. Any treatments given to the child prior to arrival or after discharge from the enrollment health facility were excluded. To reduce the risk of recall bias, children had a diarrhea diary completed by their parent/caregiver for 14 days following their visit to the health facility.

### Definitions

WHO diarrhea management guidelines indicate that zinc should be offered to each child presenting with diarrhea, and antibiotics, specifically azithromycin, ceftriaxone, or ciprofloxacin, should only be provided to children presenting with dysentery(7). This study explored two differing definitions for meeting the zinc guideline criterion: either offering the child 10 or more days of zinc or offering the child any zinc regardless of duration of prescription. Children with dysentery were considered guideline adherent if they were administered or prescribed azithromycin, ceftriaxone, ciprofloxacin, or pivmecillinam (which was considered a recommended antibiotic for dysentery in the Bangladesh site due to high prevalence of resistance to the others). Children with non-dysenteric watery diarrhea were considered guideline adherent if they were not administered nor prescribed any of the above four antibiotics. Children with non-dysenteric watery diarrhea may have received other antibiotics besides the four recommended drugs and were still considered guideline-adherent. Any use of the four recommended antibiotics in a child with non-dysenteric watery diarrhea was considered guideline non-adherent.

Diarrhea duration was defined as the number of days when the child experienced three or more loose stools from illness onset until experiencing two diarrhea-free days. For most children, duration was determined from the caregiver-completed daily diarrhea diary. If the diary was not returned or if the episode exceeded 14 days, the end of the child’s diarrhea episode was determined by caregiver report at the four-week (preferred) or three-month follow-up visit.

### Statistical Analysis

To assess the relationship between adherence to diarrhea management guidelines for antibiotics, zinc and diarrhea duration, this study employed single and multivariable linear modelling with robust standard errors for diarrhea duration with each of the guideline-specific treatments (i.e. zinc use, antibiotic use among participants with dysentery, and antibiotic use among participants with watery diarrhea) as exposures. Models were adjusted for age, sex, highest achieved maternal education, country, and presence of wasting. All statistical tests were two-sided using a 5% significance level (alpha of 0.05). The analysis was conducted using RStudio version 4.2.2. Ethical approval was obtained prior study initiation and informed consent obtained from each parent and or legal guardian.

## Results

EFGH enrolled 9,476 children aged 6 to 35 months with diarrhea across the seven EFGH sites, with 9,397 of these children having complete records, and thus included in this analysis. The majority (63.3%) of participants were under 18 months of age with minimal variation across sites; 54.4% were male (Table 1). Across all sites, severe acute malnutrition (SAM) was rare (3.6%). Stunting, wasting, and underweight were present at 23.7%, 16.8%, and 21.6% respectively, with considerable variation by site. On average, caregivers sought care for their child after two days of symptomatic diarrhea, with an average total episode duration of just under five days (4.773.24). Of enrolled children, 1,214 (12.9%) presented with dysentery.

**Table 1:**
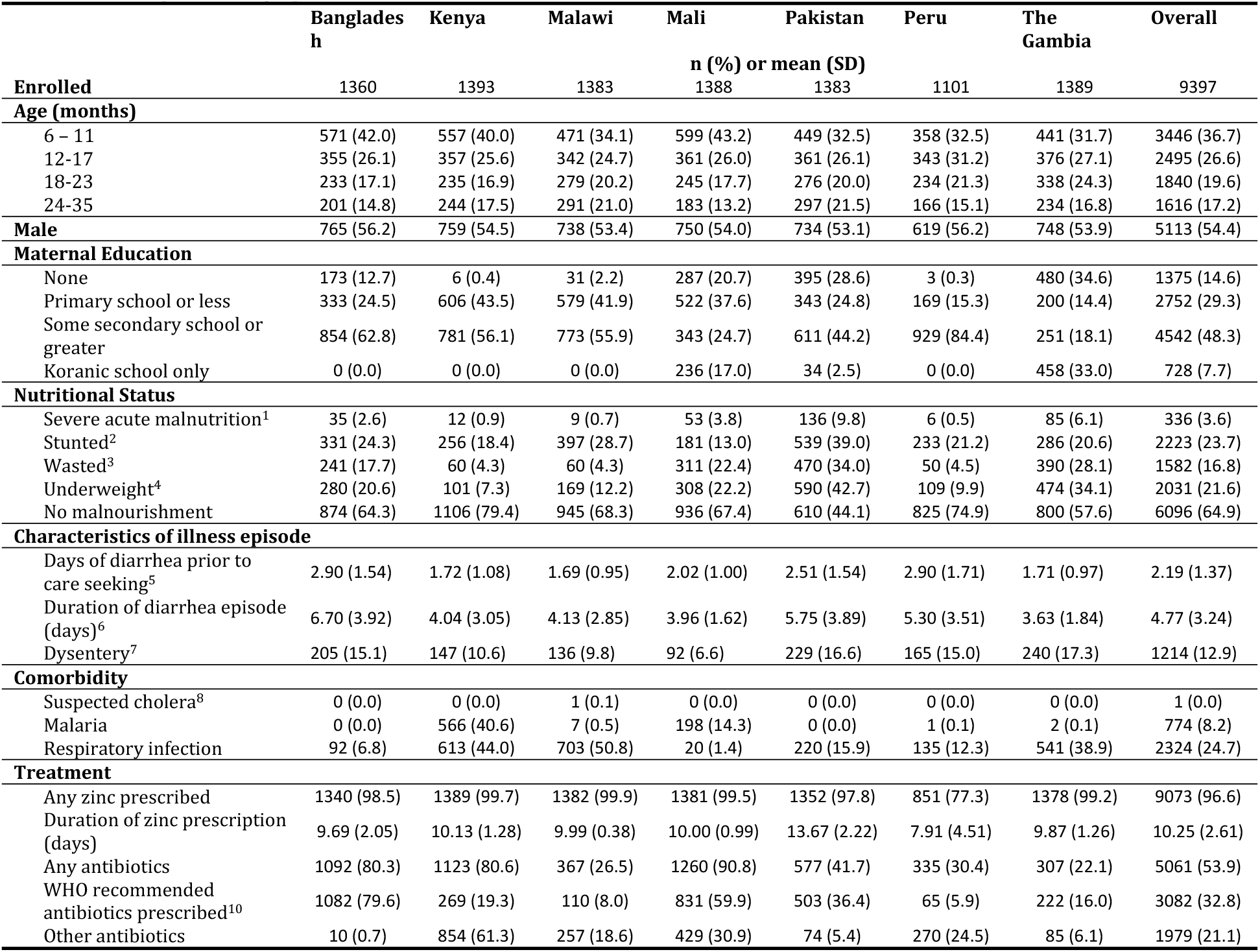
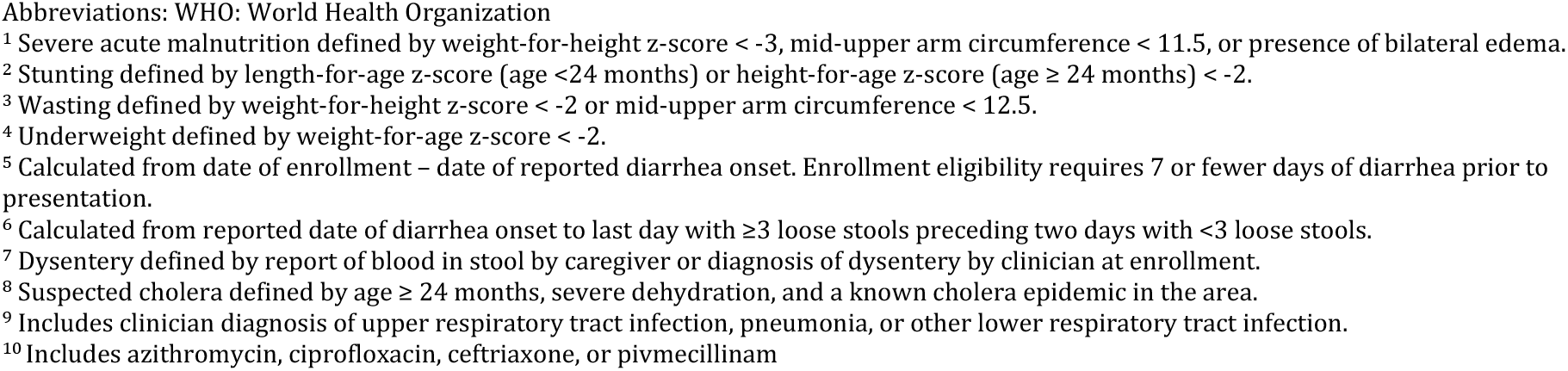
Participant demographic and illness characteristics; EFGH, 2022-2024.

|  | Bangladesh | Kenya | Malawi | Mali | Pakistan | Peru | The Gambia | Overall |
| --- | --- | --- | --- | --- | --- | --- | --- | --- |
|  | n (%) or mean (SD) |  |  |  |  |  |  |  |
| **Enrolled** | 1360 | 1393 | 1383 | 1388 | 1383 | 1101 | 1389 | 9397 |
| **Age (months)** |  |  |  |  |  |  |  |  |
| 6 – 11 | 571 (42.0) | 557 (40.0) | 471 (34.1) | 599 (43.2) | 449 (32.5) | 358 (32.5) | 441 (31.7) | 3446 (36.7) |
| 12-17 | 355 (26.1) | 357 (25.6) | 342 (24.7) | 361 (26.0) | 361 (26.1) | 343 (31.2) | 376 (27.1) | 2495 (26.6) |
| 18-23 | 233 (17.1) | 235 (16.9) | 279 (20.2) | 245 (17.7) | 276 (20.0) | 234 (21.3) | 338 (24.3) | 1840 (19.6) |
| 24-35 | 201 (14.8) | 244 (17.5) | 291 (21.0) | 183 (13.2) | 297 (21.5) | 166 (15.1) | 234 (16.8) | 1616 (17.2) |
| **Male** | 765 (56.2) | 759 (54.5) | 738 (53.4) | 750 (54.0) | 734 (53.1) | 619 (56.2) | 748 (53.9) | 5113 (54.4) |
| **Maternal Education** |  |  |  |  |  |  |  |  |
| None | 173 (12.7) | 6 (0.4) | 31 (2.2) | 287 (20.7) | 395 (28.6) | 3 (0.3) | 480 (34.6) | 1375 (14.6) |
| Primary school or less | 333 (24.5) | 606 (43.5) | 579 (41.9) | 522 (37.6) | 343 (24.8) | 169 (15.3) | 200 (14.4) | 2752 (29.3) |
| Some secondary school or greater | 854 (62.8) | 781 (56.1) | 773 (55.9) | 343 (24.7) | 611 (44.2) | 929 (84.4) | 251 (18.1) | 4542 (48.3) |
| Koranic school only | 0 (0.0) | 0 (0.0) | 0 (0.0) | 236 (17.0) | 34 (2.5) | 0 (0.0) | 458 (33.0) | 728 (7.7) |
| **Nutritional Status** |  |  |  |  |  |  |  |  |
| Severe acute malnutrition<sup>1</sup> | 35 (2.6) | 12 (0.9) | 9 (0.7) | 53 (3.8) | 136 (9.8) | 6 (0.5) | 85 (6.1) | 336 (3.6) |
| Stunted<sup>2</sup> | 331 (24.3) | 256 (18.4) | 397 (28.7) | 181 (13.0) | 539 (39.0) | 233 (21.2) | 286 (20.6) | 2223 (23.7) |
| Wasted<sup>3</sup> | 241 (17.7) | 60 (4.3) | 60 (4.3) | 311 (22.4) | 470 (34.0) | 50 (4.5) | 390 (28.1) | 1582 (16.8) |
| Underweight<sup>4</sup> | 280 (20.6) | 101 (7.3) | 169 (12.2) | 308 (22.2) | 590 (42.7) | 109 (9.9) | 474 (34.1) | 2031 (21.6) |
| No malnourishment | 874 (64.3) | 1106 (79.4) | 945 (68.3) | 936 (67.4) | 610 (44.1) | 825 (74.9) | 800 (57.6) | 6096 (64.9) |
| **Characteristics of illness episode** |  |  |  |  |  |  |  |  |
| Days of diarrhea prior to care seeking<sup>5</sup> | 2.90 (1.54) | 1.72 (1.08) | 1.69 (0.95) | 2.02 (1.00) | 2.51 (1.54) | 2.90 (1.71) | 1.71 (0.97) | 2.19 (1.37) |
| Duration of diarrhea episode (days)<sup>6</sup> | 6.70 (3.92) | 4.04 (3.05) | 4.13 (2.85) | 3.96 (1.62) | 5.75 (3.89) | 5.30 (3.51) | 3.63 (1.84) | 4.77 (3.24) |
| Dysentery<sup>7</sup> | 205 (15.1) | 147 (10.6) | 136 (9.8) | 92 (6.6) | 229 (16.6) | 165 (15.0) | 240 (17.3) | 1214 (12.9) |
| **Comorbidity** |  |  |  |  |  |  |  |  |
| Suspected cholera<sup>8</sup> | 0 (0.0) | 0 (0.0) | 1 (0.1) | 0 (0.0) | 0 (0.0) | 0 (0.0) | 0 (0.0) | 1 (0.0) |
| Malaria | 0 (0.0) | 566 (40.6) | 7 (0.5) | 198 (14.3) | 0 (0.0) | 1 (0.1) | 2 (0.1) | 774 (8.2) |
| Respiratory infection | 92 (6.8) | 613 (44.0) | 703 (50.8) | 20 (1.4) | 220 (15.9) | 135 (12.3) | 541 (38.9) | 2324 (24.7) |
| **Treatment** |  |  |  |  |  |  |  |  |
| Any zinc prescribed | 1340 (98.5) | 1389 (99.7) | 1382 (99.9) | 1381 (99.5) | 1352 (97.8) | 851 (77.3) | 1378 (99.2) | 9073 (96.6) |
| Duration of zinc prescription (days) | 9.69 (2.05) | 10.13 (1.28) | 9.99 (0.38) | 10.00 (0.99) | 13.67 (2.22) | 7.91 (4.51) | 9.87 (1.26) | 10.25 (2.61) |
| Any antibiotics | 1092 (80.3) | 1123 (80.6) | 367 (26.5) | 1260 (90.8) | 577 (41.7) | 335 (30.4) | 307 (22.1) | 5061 (53.9) |
| WHO recommended antibiotics prescribed<sup>10</sup> | 1082 (79.6) | 269 (19.3) | 110 (8.0) | 831 (59.9) | 503 (36.4) | 65 (5.9) | 222 (16.0) | 3082 (32.8) |
| Other antibiotics | 10 (0.7) | 854 (61.3) | 257 (18.6) | 429 (30.9) | 74 (5.4) | 270 (24.5) | 85 (6.1) | 1979 (21.1) |

Abbreviations: WHO: World Health Organization
<sup>1</sup> Severe acute malnutrition defined by weight-for-height z-score < -3, mid-upper arm circumference < 11.5, or presence of bilateral edema.
<sup>2</sup> Stunting defined by length-for-age z-score (age <24 months) or height-for-age z-score (age ≥ 24 months) < -2.
<sup>3</sup> Wasting defined by weight-for-height z-score < -2 or mid-upper arm circumference < 12.5.
<sup>4</sup> Underweight defined by weight-for-age z-score < -2.
<sup>5</sup> Calculated from date of enrollment – date of reported diarrhea onset. Enrollment eligibility requires 7 or fewer days of diarrhea prior to presentation.
<sup>6</sup> Calculated from reported date of diarrhea onset to last day with ≥3 loose stools preceding two days with <3 loose stools.
<sup>7</sup> Dysentery defined by report of blood in stool by caregiver or diagnosis of dysentery by clinician at enrollment.
<sup>8</sup> Suspected cholera defined by age ≥ 24 months, severe dehydration, and a known cholera epidemic in the area.
<sup>9</sup> Includes clinician diagnosis of upper respiratory tract infection, pneumonia, or other lower respiratory tract infection.
<sup>10</sup> Includes azithromycin, ciprofloxacin, ceftriaxone, or pivmecillinam

Zinc administration was high (96.6%), with over 10 days of prescribed treatment on average. Prevalence of zinc treatment was >97% across sites, with the exception of Peru, which had 77.3% administration. Among all enrolled children, 5,061 (53.9%) were offered antibiotics, of whom 3,082(32.8% of the overall population) _ received one of the four recommended medications (Table 1). Antibiotic use had considerable variability across sites, with 5.9% of all children in Peru receiving any of the recommended antibiotics and 79.6% of all children in Bangladesh receiving recommended antibiotics. Among participants presenting with dysentery, 67.6% were offered a WHO-recommended antibiotic (Table 2, figure 1). Among participants with watery diarrhea without dysentery, 72.4% were not given any of the four recommended antibiotics and were thus considered guideline-adherent. Overall, 6,302 (67.1%) of children received guideline-adherent care when combining zinc and appropriate antibiotic use (Table 2, figure 1).

**Table 2.** Children enrolled in EFGH receiving guideline-adherent care specific to zinc and antibiotic administration.

|  | Bangladesh | Kenya | Malawi | Mali | Pakistan | Peru | The Gambia | Overall |
| --- | --- | --- | --- | --- | --- | --- | --- | --- |
| **Zinc Prescribed** |  |  |  |  |  |  |  |  |
| Any zinc prescribed | 1340/1360 (98.5) | 1389/1393 (99.7) | 1382/1383 (99.9) | 1381/1388 (99.5) | 1352/1383 (97.8) | 851/1101 (77.3) | 1378/1389 (99.2) | 9073/9397 (96.6) |
| ≥10 days zinc prescribed | 1240/1360 (91.2) | 1382/1393 (99.2) | 1381/1383 (99.9) | 1381/1388 (99.5) | 1347/1383 (97.4) | 744/1101 (67.6) | 1360/1389 (97.9) | 8835/9397 (94.0) |
| **Guideline-adherent antibiotic use among those with dysentery<sup>1</sup>** |  |  |  |  |  |  |  |  |
| Guideline adherent | 182/205 (88.8) | 87/147 (59.2) | 98/136 (72.1) | 67/92 (72.8) | 159/229 (69.4) | 45/165 (27.3) | 183/240 (76.2) | 821/1214 (67.6) |
| Not adherent | 23/205 (11.2) | 60/147 (40.8) | 38/136 (27.9) | 25/92 (27.2) | 70/229 (30.6) | 120/165 (72.7) | 57/240 (23.8) | 393/1214 (32.4) |
| **Not given antibiotics with watery diarrhea<sup>2</sup>** |  |  |  |  |  |  |  |  |
| Guideline adherent | 255/1155 (22.1) | 1064/1246 (85.4) | 1235/1247 (99.0) | 532/1296 (41.0) | 810/1154 (70.2) | 916/936 (97.9) | 1110/1149 (96.6) | 5922/8183 (72.4) |
| Not adherent | 900/1155 (77.9) | 182/1246 (14.6) | 12/1247 (1.0) | 764/1296 (59.0) | 344/1154 (29.8) | 20/936 (2.1) | 39/1149 (3.4) | 2261/8183 (27.6) |
| **Guideline-adherent zinc and antibiotic administration<sup>3</sup>** |  |  |  |  |  |  |  |  |
| Among those with dysentery | 172/205 (83.9) | 86/147 (58.5) | 98/136 (72.1) | 67/92 (72.8) | 157/229 (68.6) | 31/165 (18.8) | 179/240 (74.6) | 790/1214 (65.1) |
| Among those with watery diarrhea | 183/1155 (15.8) | 1056/1246 (84.8) | 1233/1247 (98.9) | 529/1296 (40.8) | 792/1154 (68.6) | 623/936 (66.6) | 1096/1149 (95.4) | 5512/8183 (67.4) |
| Overall | 355/1360 (26.1) | 1142/1393 (82.0) | 1331/1383 (96.2) | 596/1388 (42.9) | 949/1383 (68.6) | 654/1101 (59.4) | 1275/1389 (91.8) | 6302/9397 (67.1) |

Abbreviations: WHO: World Health Organization
<sup>1</sup> Guidelines dictate that a child presenting with diarrhea and blood in stool (dysentery) should receive azithromycin, ciprofloxacin, ceftriaxone, or pivmecillinam<sup>2</sup> Guidelines dictate that a child presenting with diarrhea and no evidence of blood in stool should not be given azithromycin, ciprofloxacin, ceftriaxone, or pivmecillinam<sup>3</sup> 10+ days of zinc prescription and appropriate antibiotic prescribing given presence of blood in stool or not

**Figure 1.**
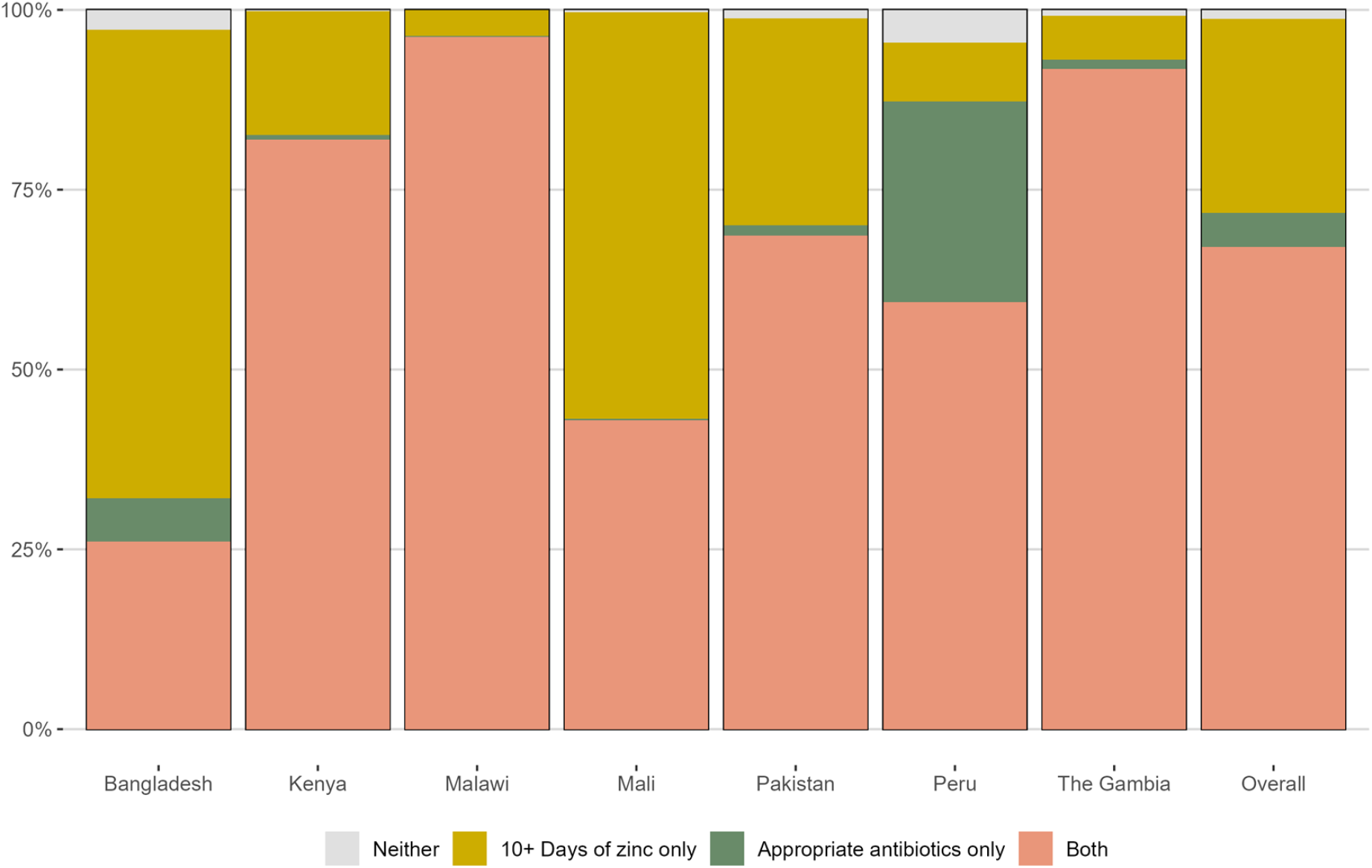
Summary of full, partial, or no guideline adherence in zinc and antibiotic treatment for diarrhea across the EFGH sites, 2022-2024

When compared to children who did not receive guideline adherent care, children with guideline adherent care had shorter diarrhea duration for all indicators except lack of antibiotic use in children with non-dysenteric diarrhea. Children who were given ≥ 10 days of zinc had on average 0.36 (95% CI: 0.03, 0.70) fewer days of diarrhea (Table 3). In children with dysentery, providing recommended antibiotics was associated with 1.08 (95% CI: 0.63, 1.53) fewer days of diarrhea compared to dysenteric diarrhea cases who did not receive the recommended antibiotics. Among all participants, receipt of zinc and appropriate antibiotic use together was associated with 0.26 (95% CI: 0.10, 0.41) fewer days of diarrhea than those with partial or non-adherent treatment (Table 3). Among cases of dysentery, receipt of both zinc and appropriate antibiotics was associated with 1.05 (95% CI: 0.59, 1.50) fewer days of diarrhea (Table 3).

**Table 3.**
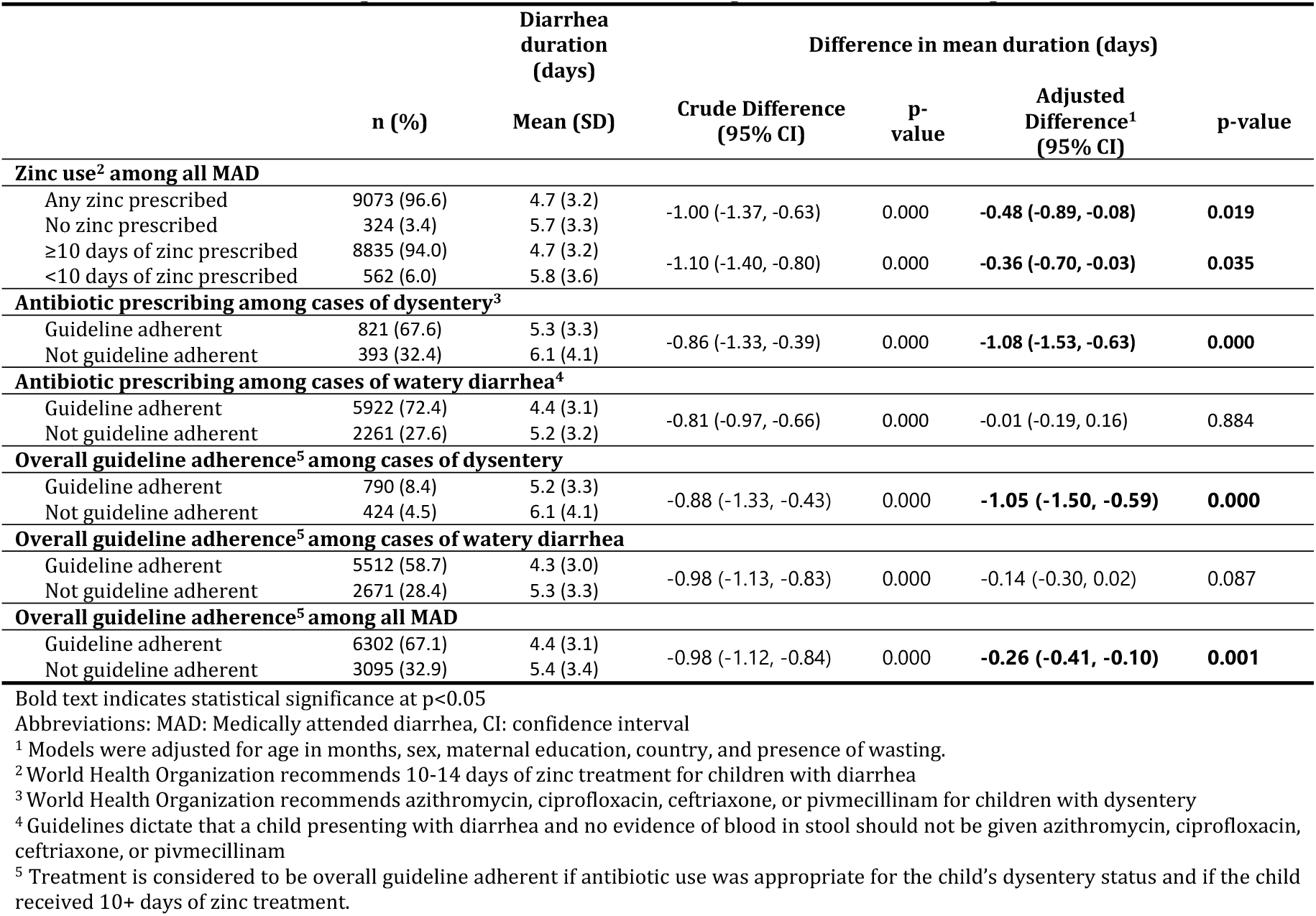
Diarrhea duration among those who did and did not receive guideline-adherent management; EFGH, 2022-2024.

## Discussion

Despite the definition of clear case management guidelines nearly 20 years ago for antibiotic use in dysentery and acute diarrhea(4) and recommendation regarding zinc(15) and reinforcement of adherence to guidelines at the study sites, just over two-thirds of participants received the recommended therapy for acute diarrhea and dysentery. Despite this, our findings reveal that adherence to WHO treatment guidelines is associated with shortened diarrhea duration in children aged 6-35 months. The majority of participants in our study were prescribed zinc for more than ten days, with only Peru having <97% zinc administration. The majority of participants received complete guideline adherent care; this finding may be explained by the involvement of study staff in case-management across sites. In a cross-sectional study from Nigeria, maternal age 30-39, when compared to maternal age 20-29 and >40, was found to be associated with increased knowledge of diarrhea management (including zinc administration for 10 days or more) (12). In all children with diarrhea, adherence to the zinc guideline was associated with reductions in duration of illness(12). Our results are similar to a meta-analysis by Ali et al. in 2024 which suggested that the duration of diarrhea decreased significantly in the zinc-treated group as compared to the placebo group (3). Two-thirds of EFGH participants with dysentery received WHO-recommended antibiotics, with the lowest rates of antibiotic prescription in Peru and the highest in Bangladesh. Similar results were reported in the Vaccine Impact on Diarrhea in Africa (VIDA) study, where 62.3% of participants with bloody diarrhea and 11.6% of participants with watery diarrhea receiving WHO recommended antibiotics (16). Our results demonstrate that receipt of zinc and appropriate antibiotics among dysenteric patients was associated with fewer days of diarrhea than those with partial or non-adherent treatment.

Antibiotic use is generally not indicated in acute watery diarrhea due to its typically self-limiting nature and risk of antimicrobial resistance, often resolving spontaneously in less than three days (17). Among participants with watery diarrhea without dysentery in EFGH, most were not given WHO-recommended antibiotics and were thus considered guideline-adherent. This guideline-adherence was not associated with reduction in diarrhea duration, likely suggesting limited benefit to illness duration with the prescribing of antibiotics when not indicated. In contrast, children with dysentery provided WHO-recommended antibiotics had significantly fewer days of diarrhea than those who did not receive antibiotics, suggesting notable improvement to illness duration when antibiotics were properly used.

Our analysis has some limitations. First, since our population is composed of children aged 6-35 months in specific settings who sought care for their illness, thus findings cannot be generalized to all children or to children with non-medically attended diarrhea. Second, EFGH provided antibiotics and zinc to prevent shortages, making it possible to have such high proportions of guideline adherent care which may not be representative of real-world conditions. Lastly, these results are from a research setting with funding support and oversight, so the generalizability to purely programmatic settings is uncertain. It is reasonable to hypothesize that settings outside of a study would have lower adherence due to increased medication stockouts and would require increased emphasis on clinical management guidelines compared to a settings under a funded international study. Nonetheless, the benefits to diarrhea duration from guideline-adherent care as demonstrated in this study would be expected to persist under real-world clinical conditions and underscore the importance of clinician training and consistent medication availability in clinical settings to promote guideline-adherent care and make it accessible for children with diarrhea. A major strength of this study is high data quality. Collected medical data was highly detailed and infilled directly by the treating clinician so we have high confidence in the accuracy of zinc and antibiotic prescribing in EFGH. Further, follow-up retention was high (>97%) with more than 90% of children having returned a completed diarrhea diary resulting in high-accuracy calculation of diarrhea duration.

## Conclusion

We found evidence that adherence to zinc and antibiotic treatment guidelines, particularly among children with dysentery, for medically attended diarrhea in children is associated with shorter diarrhea duration. We show that zinc use was very high among children presenting to care with diarrhea and that antibiotic use was appropriate in the majority of cases. Two-thirds of children with dysentery received the recommended antibiotics, and nearly three-fourths of children with non-dysenteric watery diarrhea were not given inappropriate antibiotics. These findings are important for regional quality improvement, education, and policy implementation efforts.

## Data Availability

All the data will be provided by the EFGH (Enteric for Global Health) consortium.

## Acknowledgements

We acknowledge All EFGH participants and families, clinical and study staff, Sonia Rao and Beth Barr for their facilitations.

## Funding

This research was supported by the Bill & Melinda Gates Foundation [INV-031791].

## Notes

### Competing Interest Statement

The authors have declared no competing interest.

### Funding Statement

This work was funded by the Gates Foundation (award numbers INV-031791, INV-045988, INV-062665, INV-076498). The funding organization had no role in the study design, data collection, analysis, decision to publish, or preparation of the manuscript.

### Author Declarations

This study was conducted according to Good Clinical Practice (GCP), including Good Clinical Laboratory Practice (GCLP), the Declaration of Helsinki, IRB and local rules and regulations specific to each EFGH country. This protocol was subject to ethical approval from the Institutional Review Boards (IRBs) at each EFGH site. Bangladesh: The Institutional Review Board of International Centre for Diarrhoeal Disease Research, Bangladesh gave ethical approval for this work [Approval #: PR-21114] Kenya: The KEMRI Scientific and Ethics Review Unit of the Kenya Medical Research Institute gave ethical approval for this work [Approval #: PROTOCOL NO. KEMRI/SERU/CGHR/403/4362]. Kenya Medical Research Institute, has been licensed by The National Commission for Science, Technology and Innovation to conduct research as per the provision of the Science, Technology and Innovation Act, 2013 (Rev.2014) in Siaya on the topic: ENTERICS FOR GLOBAL HEALTH: SHIGELLA SURVEILLANCE STUDY (EFGH) for the period ending : 27/January/2024 [License No: NACOSTI/P/22/15379 & NACOSTI/P/23/23350]. Malawi: The College of Medicine Research Ethics Committee of Kamuzu University of Health Sciences gave ethical approval for this work [Approval #’s: P.10/21/3437]. The Central University Research Ethics Committee D of the University of Liverpool gave ethical approval for this work [Approval #: 10596]. Mali: Universite Des Sciences, Des Techniques et des Technologies de Bamako gave ethical approval for this work [Approval #’s: 00000918, 00000964, 0000091, 00000189, 00000440, 0000022, 00000383, 00000279]. The University of Maryland, Baltimore Institutional Review Board gave ethical approval for this work [Approval #’s: HP-00098210, HM-HP-00098210-1, HM-HP-00098210-2, HM-HP-00098210-3]. Pakistan: The Ethics Review Committee of The Aga Khan University gave ethical approval for this work [Approval #’s: 2021-6932-19680, 2022-6932-21888, 2022-6932-23332, 2022-6932-23399, 2023-6932-24228, 2023-6932-25760]. The Pakistan National Institutes of Health, Health Research Institute, National Bioethics Committee gave ethical approval for this work [Approval #’s: No.4-87/NBC-746/22/1556, No.4-87/NBC-746/22/160, No.4-87/NBC-746-Exten/23/1577]. Peru: Comite Institucional de Etica en Investigacion de la Asociacion Benefica Prisma gave ethical approval for this work [Approval #’s: CE0043.22, CE0669.23, CE0674.22, CE0513.23, CE0669.23]. The Gambia: The Observational / Interventions Research Ethics Committee of the London School of Hygiene & Tropical Medicine gave ethical approval for this work [Approval #: 26515]. U.S. Coordination: The Human Subjects Divisions of The University of Washington, Seattle, USA, The University of Virginia, Charlottesville, USA, and Emory University, Atlanta, USA, determined that the activities of the coordinating bodies do not constitute human subjects research as defined by federal regulations. Therefore, review and approval by these IRBs was not required.

